# Modeling Cerebral Hemodynamics Using BOLD Magnetic Resonance Imaging and its Application in Mild Cognitive Impairment

**DOI:** 10.1101/2020.02.23.20018846

**Authors:** B.C. Henley, M.O. Okafor, I. Hajjar

## Abstract

**Objective:** This study develops a procedure and related analytical methods for deriving indices of cerebral hemodynamics in the magnetic resonance imaging (MRI) setting using resting state recordings of systemic blood pressure, pulse rate, and end-tidal CO_2_ synchronized with the MRI image acquisitions of blood oxygenation level dependent (BOLD) data as a measure of cerebral perfusion.

**Methods:** We employed the concept of Principal Dynamic Modes (PDM) to model the effect of three determinants of cerebral perfusion: mean arterial blood pressure (MABP), end-tidal CO_2_ (PETCO_2_), and pulse rate (PR). The relation between these signals and the BOLD signal were used respectively to quantify cerebral autoregulation (CA), CO_2_ vasoreactivity (CVR), and pulse rate reactivity (PRR).

**Results:** Hemodynamic indices were obtained from 129 participants with normal cognition (NC) and mild cognitive impairment (MCI). CA was reduced in MCI compared to NC in the parietal lobe, CVR was reduced in MCI in the occipital and temporal lobes, and PRR was reduced in the frontal, parietal, occipital and temporal lobes. Reduced CVR and PRR were associated with worse cognitive scores including memory and executive function.

**Conclusion:** Employed acquisition and analysis of MRI hemodynamic identified cerebral hemodynamic alterations in MCI, related to PR and ETCO2 changes.

**Significance:** This modeling approach may offer a novel way to clinically assess cerebral hemodynamics during MRI.

## I. INTRODUCTION

Impaired cerebrovascular regulation is an important factor in the pathogenesis of cognitive impairment and neurodegenerative illnesses such as Alzheimer’s disease (AD) [1-5]. Cerebral autoregulation (CA) and cerebral vasomotor reactivity to CO_2_ (CVR) are two fundamental measures of cerebral hemodynamics that are quantified by estimating the effects of changes in systemic arterial blood pressure and arterial CO_2_ tension upon changes in cerebral blood flow, respectively. These are traditionally measured using beat-to-beat changes in mean arterial blood pressure (MABP), arterial partial pressure of CO_2_ reflected by end-tidal CO_2_ (PETCO_2_), and cerebral blood flow velocity using transcranial Doppler (TCD) [6-8]. Mathematical models have been employed to study the relation between time-series recordings of MABP and PETCO_2_ which are viewed as “inputs” and cerebral blood flow (CBF) which is viewed as an “output” to a theoretical “cerebrovascular system” defined by the input-output data [9-11]. Under this model, the simultaneous fluctuations in the time-series recordings of hemodynamic data from an individual participant provide a means to derive subject-specific indices of cerebral hemodynamics. In this context, a body of studies have concluded that alterations in cerebral hemodynamics are associated with cognitive decline [12-15].

A key limitation of TCD studies is the low spatial resolution, since it is typically measured at the middle or anterior cerebral artery. This may veil potentially useful clinical information about cerebral hemodynamics in smaller regions or at higher resolutions. For this reason, interest has begun to shift towards magnetic resonance imaging (fMRI) as an alternative approach for assessing cerebral perfusion regulation and hemodynamics [16-18]. Using CO_2_challenge by increasing inhaled air CO_2_ composition to 5-8%, CVR measured using blood oxygen level dependent (BOLD) MRI has been shown to be reduced in AD [19]. However, CO_2_ challenge in an MRI suite can be cumbersome. Hence, obtaining CVR without an external increase in CO_2_ may be more convenient.

The view that the effects of cardiorespiratory changes in the BOLD signal are useful in estimating cerebral hemodynamics is a departure from the traditional view of these signals as “physiological interferences” that confound interpretation of BOLD MRI. Since the spectral bandwidth of spontaneous BOLD fluctuations due to neuronal activity overlaps with that of cardiorespiratory fluctuations, they are typically filtered-out during BOLD signal analyses [20-22]. Although removing this interference might be appropriate to estimate neuronal activation in BOLD MRI, the rationale of this study is that these fluctuations may offer a way to assess hemodynamics in the MRI setting. Hence, we developed a modeling approach to estimate cerebral hemodynamics that leverage these data. Although using BOLD MRI for deriving hemodynamics has been tried previously, our approach has a number of innovative aspects. It uses the normal respiratory fluctuations in PETCO_2_ rather than an external CO_2_ challenge. It incorporates continuous blood pressure measurement concurrent with the BOLD signal acquisition. It also models the contribution of pulse rate (PR) and MABP to CBF during BOLD MRI acquisition, an area that has been rarely focused on in MRI settings.

We incorporated the concept of Principal Dynamic Modes (PDM) to model the BOLD response to PR, MABP and PETCO_2_ fluctuations using six minutes of resting state time series data during normal breathing. The PDMs represent the basic information in the dynamic relationship between PR, PETCO_2_ and MABP (the three inputs) and the BOLD signal (the output). This work assumes that the resulting three-input model may be used to calculate a measure of CA from the MABP input and CVR from the PETCO_2_ input. BOLD signal variability reflects, in part, changes in CBF driven by local neural activity as well as neurogenic, myogenic and other mechanisms related to perfusion pressure and pH [4, 23]. TCD studies suggest that most of the systemic effects of MABP on CBF or CA occur in the low-frequency range (below 0.1 Hz) [11, 12, 24]. On the assumption that fluctuations within the same frequency range in the BOLD signal are also related to MABP, we derived CA from the resting state fMRI. We further extended our analysis by modeling the contribution of PR to CBF regulation, providing a measure pulse rate reactivity (PRR). Our aim in this report is to describe the set-up, related procedures, and analytical methods for deriving these cerebral hemodynamic indices in the MRI setting as well as investigate their alterations in mild cognitive impairment (MCI), a prodromal stage of AD dementia.

## II. METHODS

### Participant Summary

Data from 129 Brain, Stress, Hypertension, and Aging Research Program (BSHARP) participants with either normal cognition (NC) or MCI were analyzed in this study. Inclusion criteria for MCI included subjective memory concern, abnormal cognitive function determined by the Logical Memory subscale (Delayed Paragraph Recall, Paragraph A only) from the Wechsler Memory Scale-Revised (<11 for 16+ years of education, <9 for 8-15 years of education, <6 for <7 years of education), a Montreal Cognitive Assessment (MoCA) score <26, both a clinical dementia rating (CDR) global score and memory box score = 0.5, and a functional assessment questionnaire (FAQ) <9 [25]. Inclusion criteria for NC included MoCA≥26, CDR=0, and normal Logical Memory subscale (<12 for 16+ years of education, >8 for 8-15 years of education, >5 for <7 years of education). This study was approved by the Emory IRB and each participant provided a written informed consent.

### MRI acquisition

A 50 to 60-minute MRI scan was performed on each participant in supine position using a 3T scanner (Magnetom Prisma; Siemens, Erlangen, Germany). 3D T1-weighted images were acquired using a rapid gradient-echo imaging sequence: 170 measurements with repetition time (TR) = 2500 ms (7-min record); voxel size = 3.0×3.0×3.0 mm; field of view = 220 mm; 48 transversal slices with thickness = 3.0 mm; echo time (TE) = 27 ms; flip angle = 90 degrees. This analysis uses the last six of the seven-minute resting state session of the MRI protocol. Participants underwent an additional CO_2_ challenge during the same scan session as the resting state protocol. This challenge included two phases: a normocapnic phase where each participant breathed room air followed by two minutes of exposure to hypercapnia (8% CO_2_ gas) via facemask. These data were used to derive CVR using the traditional method of increasing inhaled CO_2_ enriched air and was calculated as the percent change in BOLD divided by the change in PETCO_2_ from the average of the last minute of normocapnia to the last minute of hypercapnia during the CO_2_ exposure. Participants who failed to experience an increase in PETCO_2_ of more than 1.00 mmHg from the normocapnia to hypercapnia were not included in the analysis. Figure 1 illustrates the setup of the data acquisition system at rest and during the CO_2_ challenge in the MRI suite.

**Figure 1.**
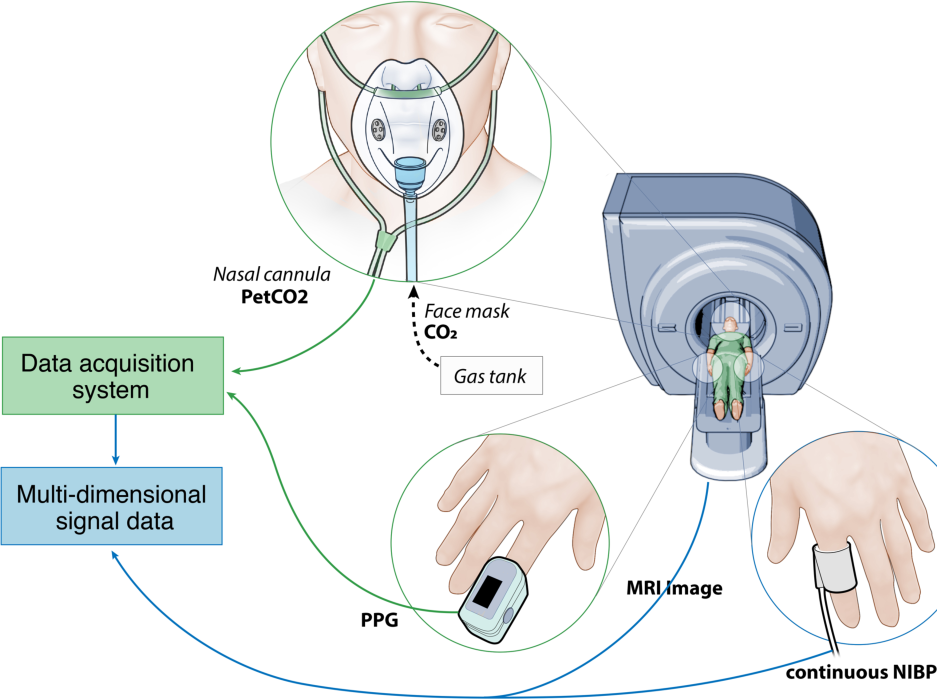
Set-up of the hemodynamic data acquisition system in the MRI suite. PETCO_2_: Partial pressure of CO_2_; NIBP: Non-invasive blood pressure; PPG: photoplethysmography.

### Cardiorespiratory signal acquisition

Simultaneously, the cardiorespiratory signals were acquired through the BIOPAC Systems hardware (MP 150). Continuous blood pressure was measured using a finger cuff placed on the thumb and connected to CareTaker© system (CT 4.5.0.12) [26]. All signals were monitored in AcqKnowledge© software (4.4.0) and analyzed using MATLAB (R2019a). Photoplethysmography was acquired using a BIOPAC BioNomadix pulse transducer. PR was computed by taking the inverse of the time interval between each systolic peak in the pulse waveform and multiplying the result by 60 to achieve the units of beats per minute (BPM). Using the systolic (SBP) and diastolic (DBP) points in the blood pressure waveform, MABP was calculated as MABP = (SBP + 2DBP)/3. PETCO_2_ was defined as the value of the %CO_2_ signal at the end of a breath and was scaled by 7.13 to convert from %CO_2_ to mmHg.

### MRI image analysis

Statistical Parametric Mapping (SPM) was used to perform image realignment and Gaussian spatial smoothing. A brain mask was used to remove voxels not corresponding to the brain. Next, the T1-weighted images were co-registered with the mean of the resting state images. The co-registered images were then normalized to Montreal Neurological Institute (MNI) template. In the MNI space, the whole brain and frontal, parietal, occipital and temporal lobes were obtained via segmentation and transformed to each participant individually. The whole brain and regional BOLD signals were finally extracted from these images. The obtained MABP, PR and ETCO_2_ signals were down-sampled to match the BOLD TR (2.5 sec), yielding three cardiorespiratory datapoints for each BOLD datapoint in the time series. All signals were then low-pass filtered at 0.1 Hz to protect the analysis from possible effects of aliasing, since the sampling rate of the BOLD signal (1/TR = 0.4 Hz) implies a Nyquist rate of 0.2 Hz, below the respiratory (∼0.3 Hz) and cardiac (∼1.0 Hz) cycle [21, 22, 27]. The voxel-wise resting state BOLD data was normalized by dividing the data by the standard deviation of the average CSF signal. The BOLD data obtained during CO_2_ challenge was divided by the last 60 seconds of the average CSF signal before the onset of CO_2_. An example of a preprocessed resting-state BOLD and cardiorespiratory data from one participant is presented in Figure 2.

**Figure 2.**
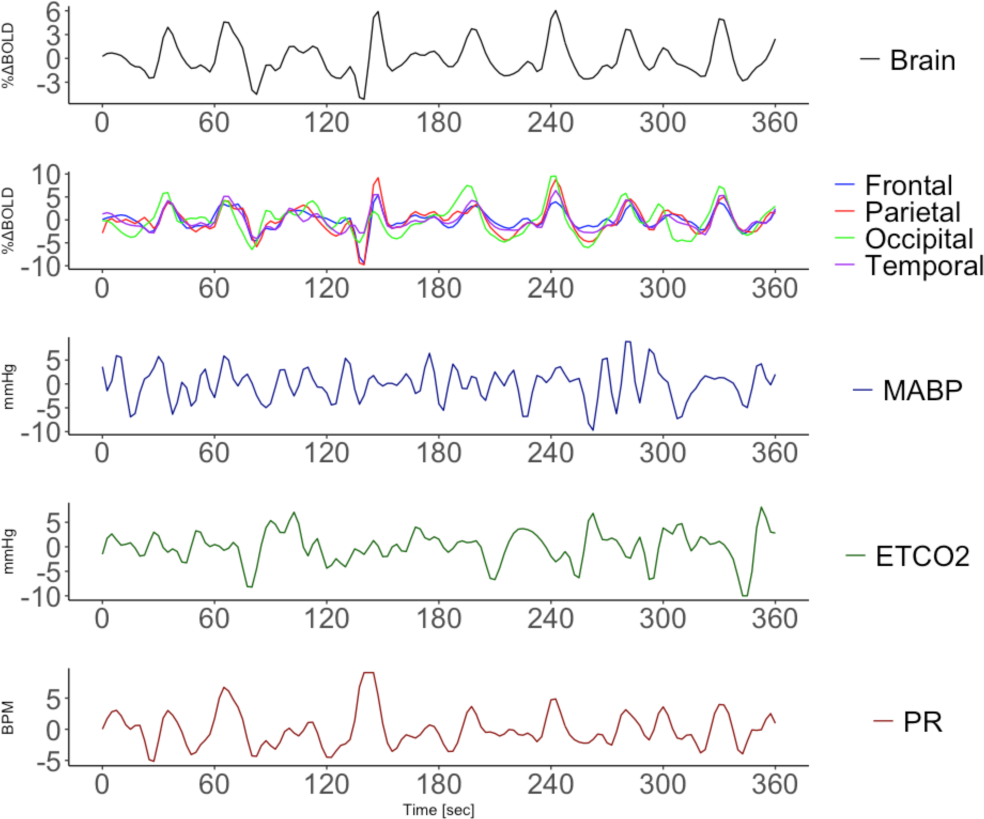
An example of 6 minutes of preprocessed time-series data recorded from a single participant, representing fluctuations in BOLD (top two panels: whole brain and lobar regions) and TR-averaged fluctuations in MABP (third panel), ETCO2 (fourth panel), and PR (bottom panel: beats per minute, BPM).

### Modeling and Statistical Methods

The dynamic effects of fluctuations in MABP, ETCO_2_ and PR upon BOLD intensity were estimated and analyzed using the PDM modeling concept. The PDM method has been used successfully to model cerebral hemodynamics using TCD and near-infrared spectroscopy (NIRS) in previous studies [24]. The PDM method is based on the Volterra model, a generalized mathematical representation of the input-output relation of a dynamic system. The pattern of this relation is defined by the Volterra kernels, which at a given time, quantifies the relative contribution of the present and past values of the input signal in generating the present value of the output signal. In this work, we defined a novel voxel-wise hemodynamic mapping where each voxel of the brain image (≈ 196,900 *per participant*) is treated as an output with MABP, PR, PETCO_2_ defined as the three inputs. As illustrated by the block diagram in Figure 3, the input-to-output transformation performed by the PDM model consists of the cascade of dynamic and static operations. The dynamic transformation is defined by the convolution of the inputs with their respective PDM. Then, the output of each convolution is separately received by associating gains that determining the relative contribution of each PDM output to the BOLD value at each voxel. The voxel components are then summed together (along with a constant offset) to form the model output prediction at that voxel.

**Figure 3.**
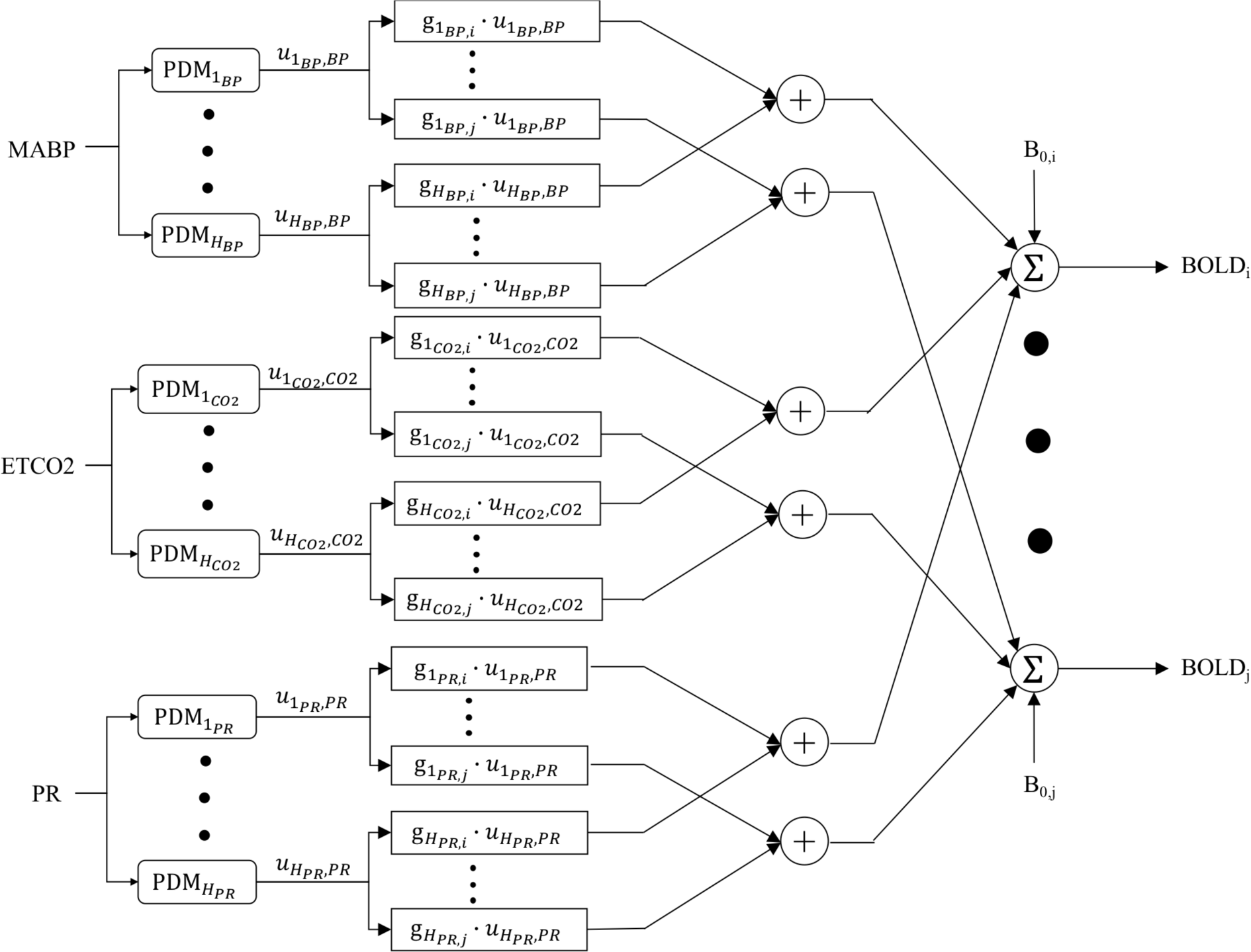
Block diagram of the PDM model for three inputs MABP, ETCO_2_ and PR, and voxel-wise percent BOLD change output. Each PDM receives their respective input, x(t), by convolution to generate the PDM output, 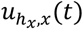. Each PDM output is scaled by their respective gains, 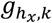. The predicted voxel-wise output, *BOLD*_*k*_(t), is the sum all components, 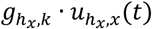 and a constant offset *BOLD*_0,*k*_.

The PDMs of the three inputs were obtained separately by first estimating the kernels using the Laguerre Expansion Technique (LET) [28] from the NC group data. Background information for this approach is provided in the Appendix. The LET model is specified by three parameters: the number (L) of orthonormal Laguerre basis functions, the relaxation parameter (α) that determines the memory of the system, and the model order (Q) which defines the degree of interaction of the input epoch. For the purpose of finding the appropriate value for these parameters, we assumed a linear LET model (Q = 1) and searched for the L and α that minimized the normalized mean square error (NMSE) for each input individually with the whole brain BOLD signal taken as the single output. Using the obtained L and α, the voxel-wise kernels were estimated using a three-input model. Singular value decomposition (SVD) was applied to the matrix containing the voxel-wise kernels of each input individually for all participants in the NC group. The smallest set of singular vectors required to explain at least 90% of the variance in the kernel matrix was selected as the PDMs for that input. The PDM gains were estimated for each participant using their input-output data (Appendix). Since this study is constrained to linear analysis, the PDMs and their respective gains contain all the information required to quantify reactivity for each participant.

### Hemodynamic indices

In order to quantify the cerebrovascular hemodynamic characteristics, we computed the model-predicted voxel-wise steady-state percent change in BOLD response to a step change in MABP, ETCO_2_and PR. These quantities are referred to as BP reactivity or cerebral autoregulation (CA), CO_2_vasoreactivity (CVR), and the newly defined PR reactivity (PRR), respectively. We note that each quantity is obtained by applying a step signal at the respective input while the other three are held constant at zero. These quantities reflect *static* hemodynamic indices, since we use the steady state %ΔBOLD value at each voxel. In addition, we quantify *dynamic* reactivity by way of the estimated PDM gains which quantifies frequency-dependent hemodynamic indices. Whole brain and regional estimates are computed by averaging over all brain tissue voxels and voxels corresponding to a region in the MNI space, respectively.

To assess the significance and utility of these indices, we examined their correlation with two additional measures of hemodynamics collected on each participant: 1) CO_2_-reactivity derived from a hypercapnic challenge protocol described above and 2) the ratio of the sit-to-stand change in MABP to PR. The latter is assumed to be an indirect measure of autonomic BP regulation in part controlled by CA and baroreceptor activity [29]. We next compared the derived PDM-based indices between NC and MCI in four brain regions: frontal, temporal, parietal and occipital region. Lastly, after adjusting for potential confounders (age, race, sex, body mass index and hypertension), we examined the associations between these indices and cognitive performance on MoCA, delayed recall on the Hopkins Verbal Learning Test (HVLT), a measure of memory, and Trail Making Test (Parts B-A), a measure of executive functioning [25, 30].

## III. RESULTS

### Participants

Our sample consists of 129 participants (mean age 65, 80 female, 69 white, 60 black, 79 MCI). Table 1 provides key characteristics of the analytical sample.

**TABLE 1.** Demographic and hemodynamic characteristics of the 129 participants included in this study.

|  |  | Normal<br>n=50 | MCI<br>n=79 |  |
| --- | --- | --- | --- | --- |
| | | $\mu(\sigma)/n(\%)$ | $\mu(\sigma)/n(\%)$ | p-value |
| Age |  | 61.59<br>(6.28) | 66.36<br>(8.45) | 0.002 |
| Sex | Male | 32<br>(65.31%) | 20<br>(37.04%) | 0.008 |
|  | Female | 17<br>(34.69%) | 34<br>(62.96%) |  |
| Race | White | 27<br>(55.1%) | 28<br>(51.85%) | 0.89 |
|  | Black | 22<br>(44.9%) | 26<br>(48.15%) |  |
| Body Mass Index |  | 27.59<br>(5.05) | 30.38<br>(6.13) | 0.03 |
| Hyper-tension | No | 20<br>(41.67%) | 27<br>(50%) | 0.52 |
|  | Yes | 28<br>(58.33%) | 27<br>(50%) |  |
| MoCA |  | 26.27<br>(2.61) | 21.22<br>(3.99) | < 0.001 |
| Sit-to-Stand MABP |  | 3.26<br>(9.36) | 6.58<br>(6.81) | 0.04 |
| Sit-to-Stand PR |  | 10.02<br>(5.28) | 8.22<br>(5.7) | 0.10 |

### Model Development and performance

Following the analytical procedure described in the Methods section, L and α were identified for all three inputs using a search procedure on all participants in the NC group which yielded the following results: L = 4 and α = 0.25 for the MABP and PR inputs and L = 4 and α = 0.4 for the PETCO_2_ input. Four PDM for each input were found to be required to explain at least 90% of the variability in the three kernel matrices. Modeling performance of observed BOLD signal demonstrated good predictive performance reflected by comparable NMSE in the NC group (used for obtaining the PDM) and the MCI group (Table 2, Supplement). Figure 4 illustrates the observed and the model-predicted whole brain BOLD output for a single participant in the MCI group, indicating good predictive performance.

**Figure 4.**
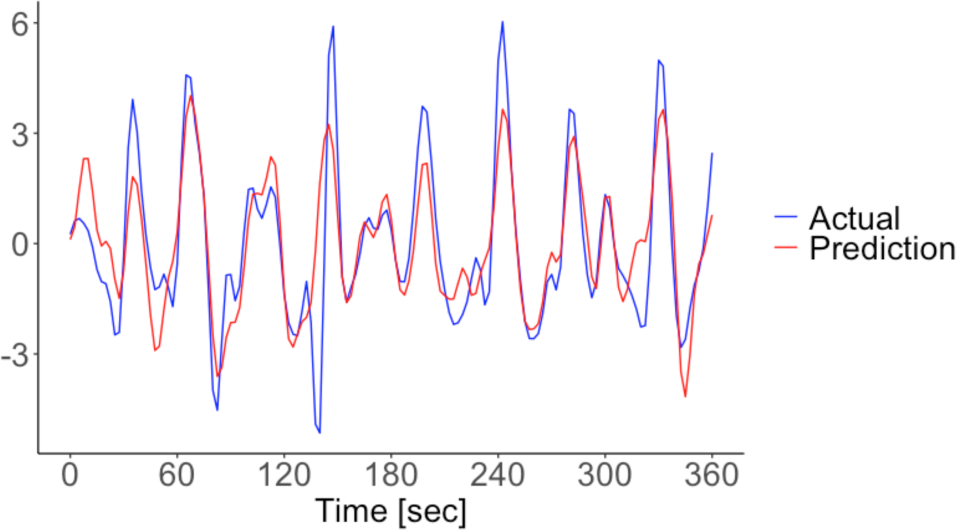
Actual and PDM-derived predicted whole brain BOLD signal for a single MCI participant showing good prediction of BOLD signal fluctuations using the PDM modeling approach.

### Dynamic characteristics of obtained PDM models

Based on our modeling criteria, we identified four PDM for each cardiorespiratory input which are illustrated in the time and frequency domains in Figure 5. Each PDM exhibited distinct resonant characteristics below 0.1 Hz, similar to that found in previous work using NIRS [24]. The first PDM for the MABP and PR had a general high-pass characteristic with a blunt peak at about 0.08 Hz for MABP and two peaks at 0.02 and 0.10 Hz for PR. The first PDM for the PETCO_2_ input exhibited a more integrative characteristic with a dominant positive phase and blunt resonant peak at about 0.07 Hz. Common among all three inputs where two PDMs that had strong resonances between 0.02 and 0.06 Hz.

**Figure 5.**
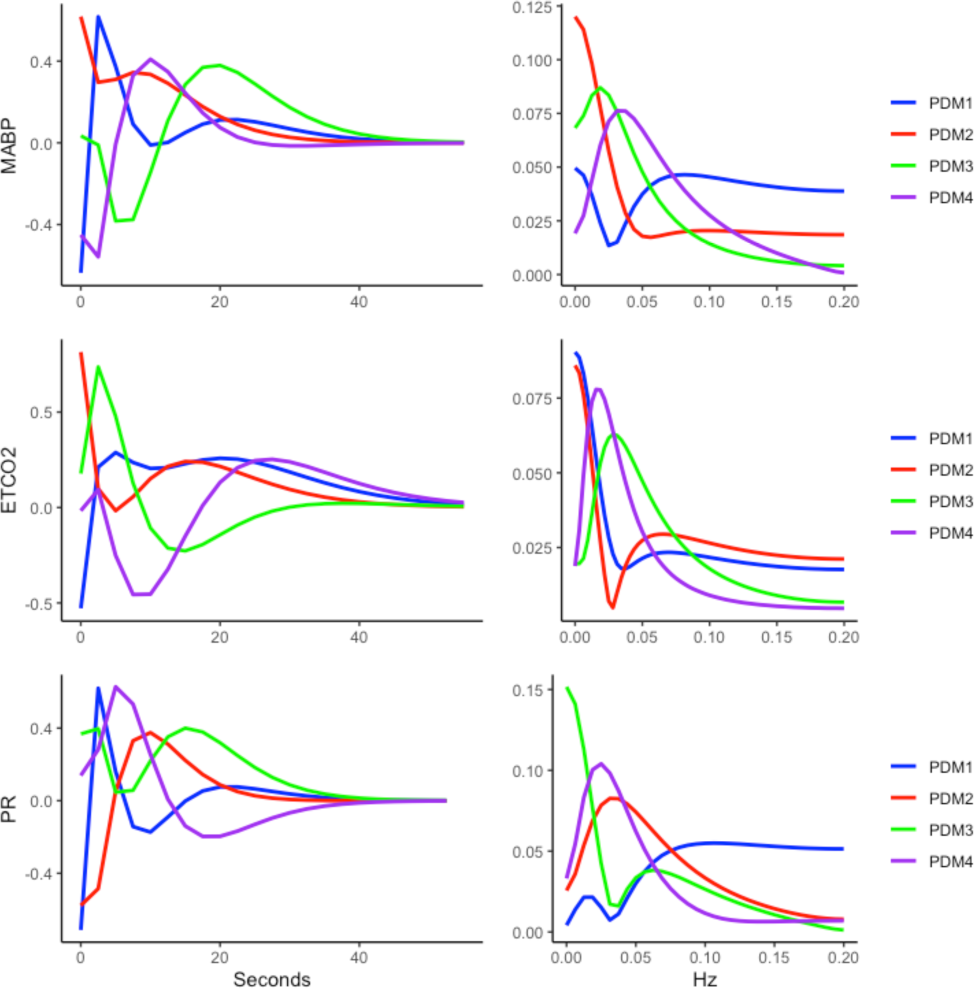
PDM for the MABP (top), ETCO_2_ (middle) and PR (bottom) inputs in the time (left) and frequency (right) domain.

Figure 6 illustrates the average whole brain step response to each input between NC and MCI, along with 95% confidence bounds. Qualitatively, there was a clear difference in the average BOLD response to the three stimuli between NC and MCI. The response to MABP appeared to exhibit the most intragroup variability, while the response to the ETCO_2_ and PR inputs showed less variability. The indices described in *Methods* were derived to test for possible statistical differences between the two groups.

**Figure 6.**
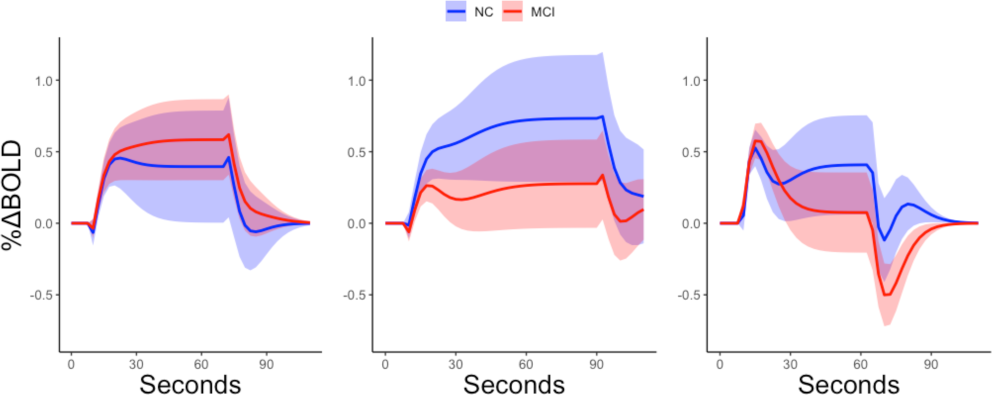
Mean (95% CI) model-predicted whole brain BOLD step response to MABP (left), ETCO_2_(middle), and PR (right) for the NC (blue) and MCI (red) groups.

### Traditional vs. PDM-based hemodynamic indices

Figure 7 illustrates the comparison between the two methods of calculating CVR and CA. There was a positive association between the two measurements of CVR (β = 0.03, p = 0.02, partial R^2^ = 0.05). There was a negative association between the ratio of sit-to-stand MABP to PR and BPR (β = −0.81, p = 0.005, partial R^2^ = 0.07).

**Figure 7.**
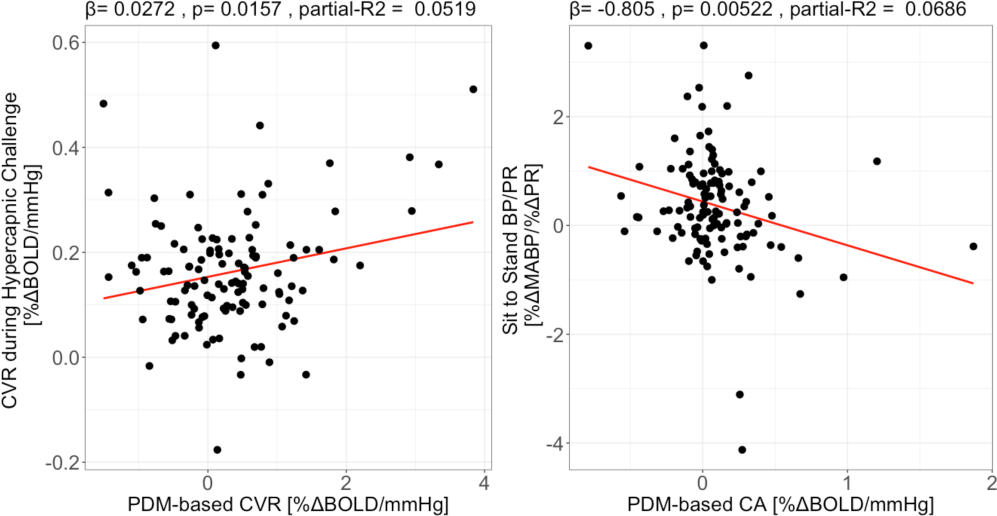
Association between a traditional measurement of CVR using hypercapnic challenge data and the PDM-derived CVR (left), and between the ratio of sit-to-stand MABP to PR and CA (right). β = slope.

### PDM-hemodynamic indices in MCI vs. NC

Figure 8 and 9 (Supplement) illustrate respectively the differences in obtained indices between NC and MCI in the four lobes and whole brain. Compared to NC, CA and the gain of the 1^st^ PDM of the MABP input was reduced in MCI in the parietal lobe. CVR was reduced in MCI in the occipital and temporal lobes, while there was no reduction in CVR using the traditional method (Table 3, Supplement). In the MCI group, the gain of the 1^st^ PDM of the ETCO_2_ input was reduced in the occipital lobe. PRR was reduced in the frontal lobe in the MCI group. The gain of the 3^rd^ PDM of the PR input was reduced in the frontal lobe, while the that of the 4^th^ PDM was reduced in whole brain and in the parietal, occipital and temporal lobes.

**Figure 8.**
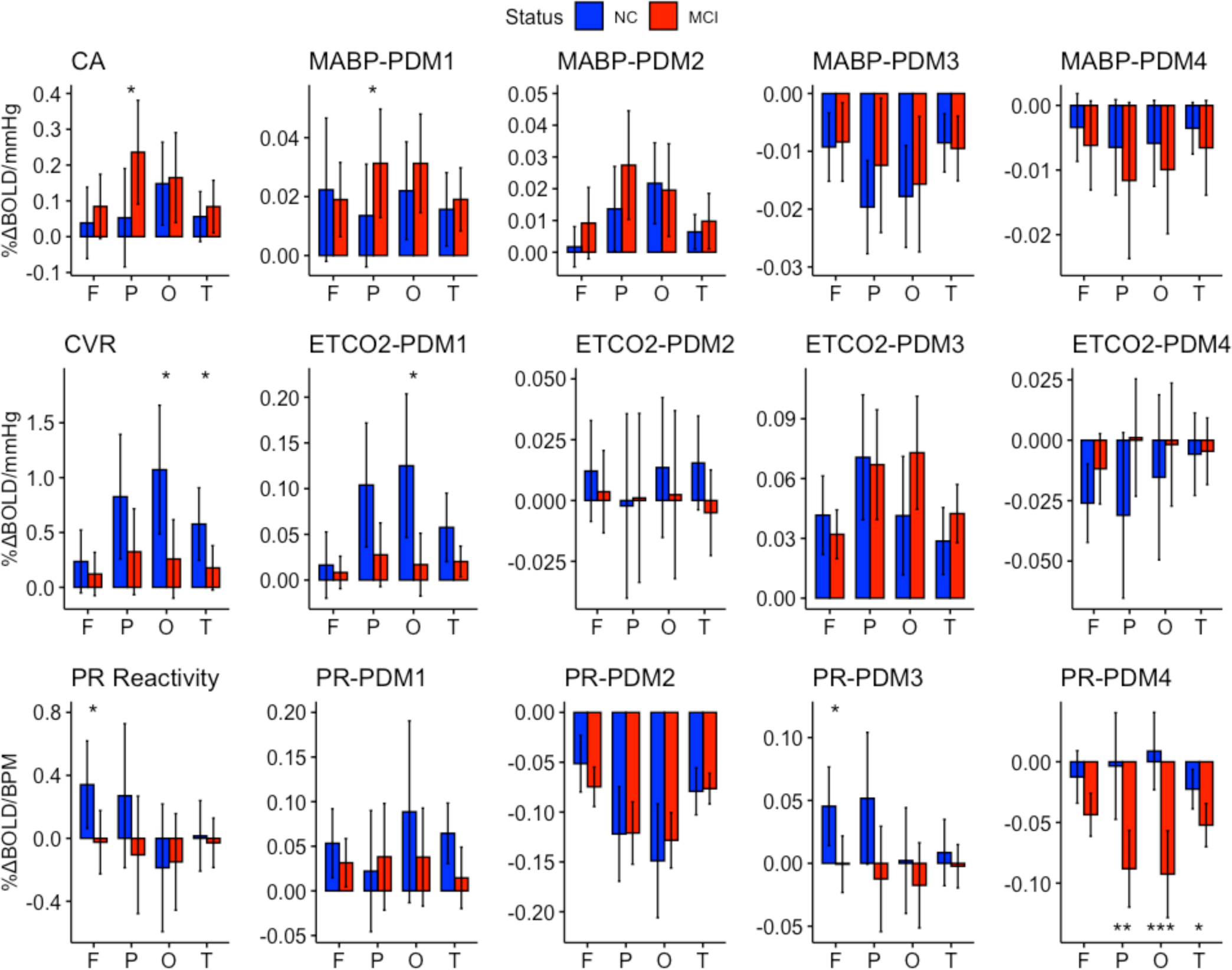
Left panel: Average CA (top), CVR (middle), and PRR (bottom) in the frontal (F), parietal (P), occipital (O) and temporal (T) lobes. Right four panels: PDM gains of the MABP (top), ETCO_2_ (middle), and PR (input) in NC (blue) and MCI (red). *: p < 0.05, ** p < 0.01, *** p < 0.001.

### Associations between cognitive scores and PDM-based hemodynamic indices

We show the association of two hemodynamic indices (CVR and PRR) that were different between the NC and MCI groups, with neuropsychological scores in Figures 10 and 11 for the four lobes and whole brain in Figure 12 (Supplement). CVR was positively associated with delayed recall on HVLT (p = 0.01) and negatively associated with Trail Making Test B-A (p = 0.02) in the occipital lobe, while both were marginal in the temporal lobe (HVLT: p = 0.07, Trails B-A: p = 0.12). The positive association of whole brain CVR with HVLT delayed recall was marginal (p = 0.092). The gain of the 1^st^ PDM of the PR input was positively associated with HVLT in the frontal (p = 0.0057), parietal (p = 0.019), occipital (p = 0.016) and temporal (p = 0.029) lobes and in Trails B-A in the frontal lobe (p = 0.048). There was a marginal association with MoCA in the frontal (p = 0.067), parietal (p = 0.098), occipital (p = 0.053) and temporal (p = 0.084) lobes. The positive association of whole brain PRR with MoCA was marginal (p = 0.092) but significant with HVLT delayed recall (p = 0.006).

**Figure 10.**
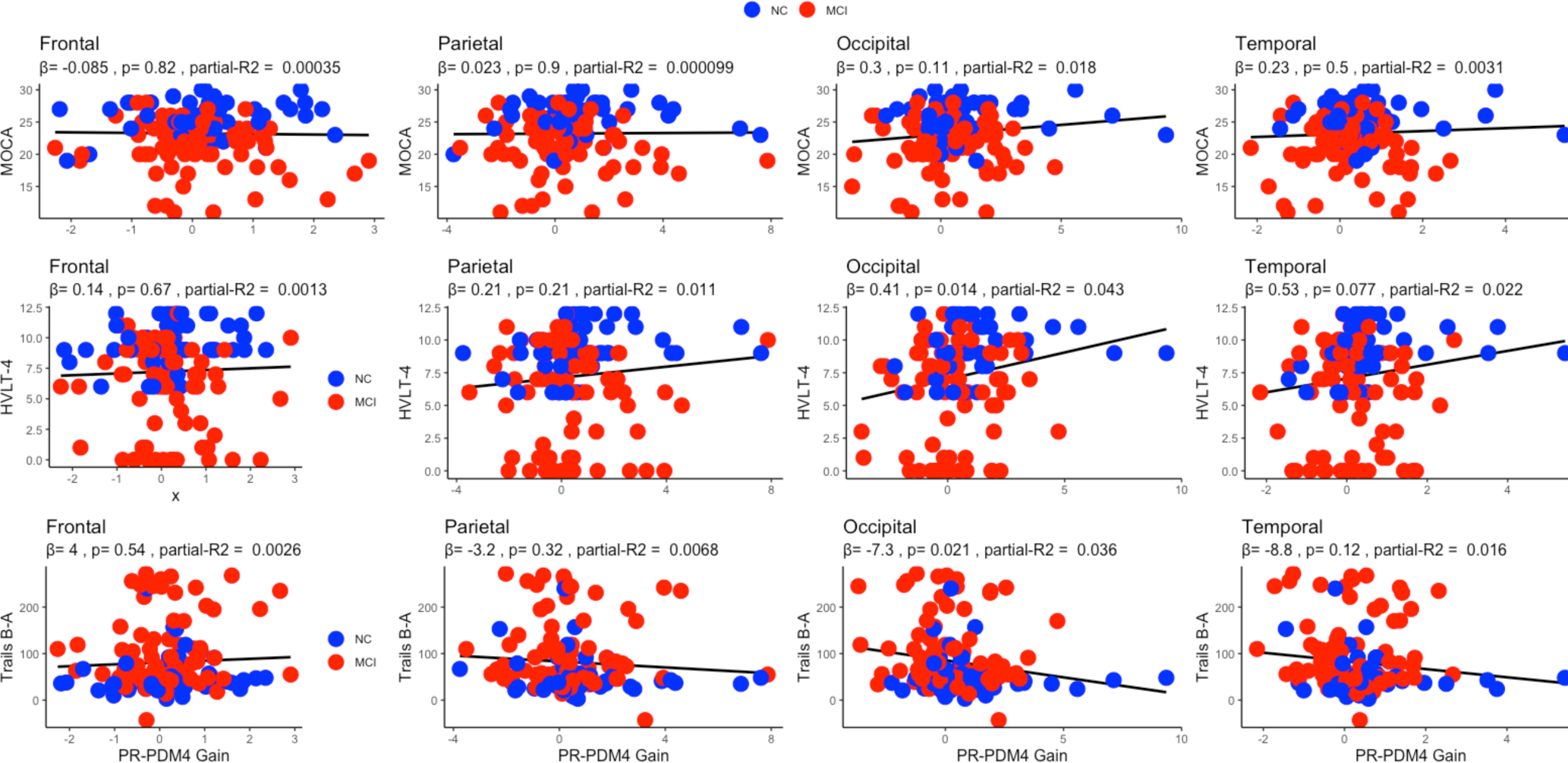
Association between the PDM-based CVR using resting state BOLD and MoCA (top), delayed recall HVLT (middle), and Trail Making test B-A (model) in the frontal (F), parietal (P), occipital (O) and temporal (T) lobes. β = slope.

**Figure 11.**
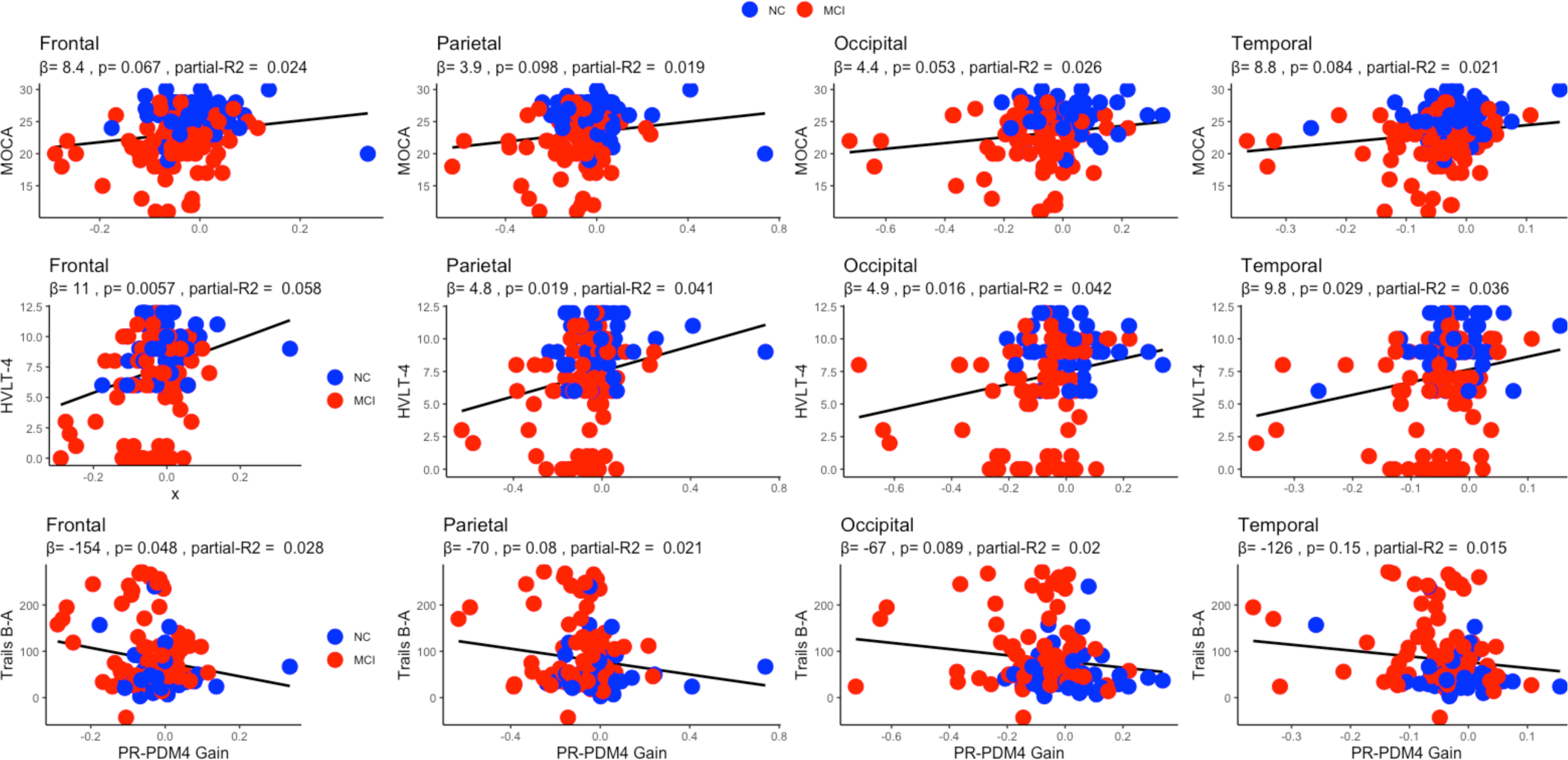
Association between the gain of the 4^th^ PDM of the PR input and MoCA (top), Delayed recall HVLT (middle), and Trails B-A (model) in the frontal (F), parietal (P), occipital (O), and temporal (T) lobes. β = slope.

## IV. Discussion

The main results of this study suggest that with moderate alterations to the MRI setup and a modeling approach to MRI data analyses, we were able to derive a set of previously described and novel indices of cerebral hemodynamics. We also discovered that PRR, an indicator of CBF response to changes in pulse rate is reduced in MCI and that this reduction is associated with cognitive performance on neuropsychological assessments. In addition, the results suggest that hemodynamic indices, especially CVR, calculated from spontaneous BOLD and cardiorespiratory fluctuations recorded during resting state fMRI using a linear input-output model has similar or even better utility than the hypercapnic protocol. The observed reduction in CVR using the PDM-based measure that was absent using the data obtained during hypercapnic challenge further suggests that measuring CVR from resting state data may be a better alternative than the traditional approach. These are consistent with our previous studies which indicate that CVR alterations at rest provide better correlates with cognition in MCI using TCD and NIRS [14, 24]. The results of this MRI study indicate that CA is not different in MCI relative to NC. Pulse rate has been shown to predict heart disease and other complications [31]. To our knowledge, however, this study is the first to demonstrate a reduction in the BOLD response to pulse rate in MCI.

One implication of the performance of the PDM-based measures in explaining differences in cognitive function relative to the traditional CVR measurements is that there may exist a more cost-effective alternative to assessing CVR in the MRI setting. This finding was also reported using TCD [15], and motivates further research. In addition, our results provide a rationale to develop and examine hypotheses about the interaction between pulse rate and cognitive function. Assuming a relatively constant stroke volume, a blunted BOLD response to a change in pulse rate may be secondary to a reduced cardiac output in MCI. We have previously reported that a state of inotropic insufficiency is present in those with MCI. This work further supports our prior findings [32].

Similar to previous studies, the estimated dynamic response of BOLD to pulse rate and CO_2_ tension mimics the putative hemodynamic response in fMRI [22, 33]. In addition, our preliminary results suggest that the dynamic BOLD response to MABP also follows this pattern. Since we measured continuous blood pressure in this study, these data provide more robust evidence for the BOLD signal-MABP association. Further, the first PDM of each input is qualitatively similar to the previously described hemodynamic waveform. A potentially insightful finding of this work is the additional PDM which do not emulate the hemodynamic response but were required to obtain sufficiently accurate adequate CBF modeling. While it is not possible to examine the physiological basis of these additional waveforms in this present study, our findings may provide a foundation to explore the possibility that these PDM may reflect other autonomic, myogenic, or yet to be discovered mechanisms.

An important limitation of our study is the high TR (2.5 sec) leading to a lower sampling rate relative to the cardiac (∼ 1 Hz) and respiratory (∼ 0.3 Hz) cycles, which is likely to lead to aliasing of the recorded BOLD time series [27]. A correction technique that relies on electrocardiogram (ECG) and respiratory phase information has been used [34]. Since ECG data was not available in our study, we used a lowpass filter with a cutoff of 0.1 Hz, which would provide a similar but not identical correction factor. Future work should examine the effect of shortening the TR on the PDM modeling results. Another important limitation of this work is the absence of the examination of the relation between the model-based hemodynamic indices and structural information (e.g. cortical thickness) and AD-related biomarkers (β-amyloid and Tau). This issue may be addressed in future work in the context of voxel-wise analysis. With these limitations in mind, the main results of this study indicate that cerebral hemodynamics can be assessed during resting state in an MRI setting. The PDM concept allowed us to measure known CA and CVR, and novel (PRR) indices of hemodynamic control, as well as perform comparisons within the brain regions for hemodynamic alterations.

## Data Availability

Data is available upon request

## V. APPENDIX

The input-output transformation performed by the single-input-single-output linear and time-invariant model is defined as

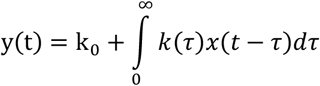

where y(t) and x(t) denote the output and input, respectively. The terms k_0_ and k(τ) are respectively the 0^th^ order kernel and first order kernel, or impulse response function, of the system. The kernels are estimated from the input-output data, which are in discrete time form. Therefore, the model above is estimated using Laguerre kernel expansion, which yields the modified Volterra model [28]:

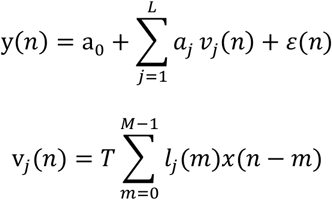

and

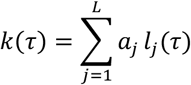

where n = t · T is the discrete time index, T is the sampling interval, M is the system memory, L is the number of Laguerre functions {*l*_*j*_} used for kernel expansion, and {a_j_} is the expansion coefficient. Estimation of the expansion coefficients involves minimizing the model prediction error *ε(n)* in the mean-square sense. This is achieved using the normalized mean-square error (NMSE):

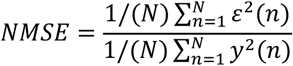

The PDM 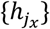 are the left singular vectors of singular value decomposition (SVD) of the matrix containing the estimated impulse response functions of input x in its columns. The number of PDM, *H*_*x*_, is defined here to be the minimum number of singular vectors required to explain 90% of the variance in the kernel matrix. This paper extends the analysis to the case of multiple outputs:

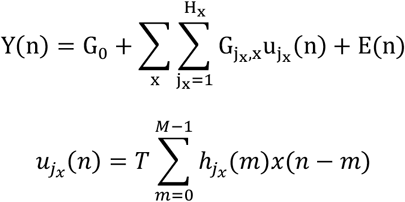

and

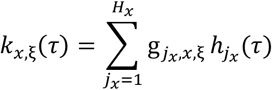

where Y(n) and E(n) are a Ξ-row vectors and Ξ is the number of output signals. The H_x_ × Ξ matrix 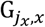 contains the PDM gains 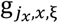 for the j_x_^th^ PDM of input x and output y_ξ_. The kernel of the ξ^*th*^ voxel, *k*_*x*, ξ_ (τ), is constructed by the PDM and their associated gains. The NMSE for the case of multiple outputs defined as:

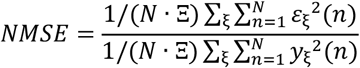

## Notes

This work was supported in part by Emory’s training grant T32NS748018 NIH grants R01AG049752, R01AG042127

### Competing Interest Statement

The authors have declared no competing interest.

### Funding Statement

This study was supported by grants
AG051633 (Dr Hajjar), AG057470-01 (Dr Hajjar), and AG042127 (Dr Hajjar), from the NIA/NIH.

